# Mendelian randomization study of spermine oxidase and cancer risk

**DOI:** 10.1101/2020.10.22.20217935

**Authors:** João Fadista, Victor Yakimov, Urmo Võsa, Christine S. Hansen, Silva Kasela, Line Skotte, Frank Geller, Julie Courraud, Tõnu Esko, Viktorija Kukuškina, Alfonso Buil, Mads Melbye, Thomas M. Werge, David M. Hougaard, Lili Milani, Jonas Bybjerg-Grauholm, Arieh S. Cohen, Bjarke Feenstra

## Abstract

**Importance:** Spermine oxidase (SMOX) catalyzes the oxidation of spermine to spermidine. Observational studies have reported SMOX as a source of induced reactive oxygen species associated with cancer, implying that inhibition of SMOX could be a target for chemoprevention.

**Objective:** To test the causality of SMOX levels with cancer risk using a Mendelian randomization analysis.

**Design, Setting, and Participants:** We performed a GWAS of spermidine/spermine ratio, from blood of 534 infants, to identify genetic variants associated with regulation of SMOX activity. In two additional data sets of 262 newborns and 508 adults, we quantified *SMOX* gene expression using RNA-sequencing and performed expression and methylation QTL lookups. We then did a Mendelian randomization analysis by testing the association between the SMOX genetic instrument and various cancer types using GWAS summary statistics.

**Main Outcomes and Measures:** Neuroblastoma, gastric, lung, breast, prostate, and colorectal cancers.

**Results:** The GWAS of spermidine/spermine ratio identified a genome-wide significant locus (P=1.34×10^−49^) explaining 32% of the variance. The lead SNP rs1741315 was also associated with *SMOX* gene expression in newborns (P=8.48×10^−28^) and adults (P=2.748×10^−8^) explaining 37% and 6% of the variance, respectively. rs1741315 was not associated with neuroblastoma (OR=0.95; 95% CI:0.88, 1.03; P=0.18), gastric (OR=0.99; 95% CI:0.95, 1.03; P=0.54), lung (OR=0.97; 95% CI:0.94, 1.00; P=0.08), breast (OR=0.99; 95% CI:0.96, 1.02; P=0.47), prostate (OR=0.98; 95% CI:0.96, 1.00; P=0.05) nor colorectal cancer (OR=1.03; 95% CI:0.99, 1.07; P=0.10). A PheWAS of rs1741315 did not reveal any associations with risk factors of the cancers tested.

**Conclusions and Relevance:** Genetic variation in the *SMOX* gene was strongly associated with SMOX activity in newborns, and less strongly in adults. Genetic down-regulation of SMOX was not significantly associated with lower odds of neuroblastoma, gastric, lung, breast, prostate and colorectal cancer. Further studies are needed to understand the effect of SMOX inhibition in relation to cancer risk.

**ARTICLE SUMMARY:** *Question:* Is SMOX causally associated with risk of cancer?

*Findings:* In this Mendelian randomization study, genetically lower levels of SMOX were not associated with decreased risk of neuroblastoma, gastric, lung, breast, prostate and colorectal cancer.

*Meaning:* These findings do not support a causal association between SMOX activity and risk of cancer, suggesting that ongoing efforts to identify SMOX inhibitors for chemoprevention may not be successful.

**STRENGHTS AND LIMITATIONS:**

- Previous studies which examined SMOX activity and cancer risk were susceptible to recall bias, confounding and reverse causation, none of which are concerns of this Mendelian randomization study.
- Our genetic instrument explained a sizeable proportion of the variance of SMOX activity
- We used summary statistics from the largest meta-analyses of primary cancer GWAS to date.
- Elevated SMOX levels in cancer could also be due to environmental factors not captured by genetics.
- Our genetic instrument was developed based on normal range SMOX activity data, thus additional genetic variants might play a role in aberrant expression of this enzyme.

## INTRODUCTION

Although polyamines are essential for normal cell growth and development^1,2^, dysregulation of polyamine metabolism is involved in tumorigenesis^3^, and hence recognized as a potential target for chemotherapy and chemoprevention^4^. Already in the 1960s, ornithine decarboxylase (ODC), the first rate-limiting enzyme in polyamine biosynthesis, was demonstrated to be at high levels in human cancer specimens^5^. Moreover, a genetic variant in ODC, regulating its enzymatic activity, confirmed the role of this enzyme in human colon cancer risk^6,40^, and ODC levels have been shown to be elevated in human skin, breast and prostate cancer specimens^7–9^. Other polyamine metabolic enzymes have not yet been reported to be associated with tumorigenesis in humans.

Spermine oxidase (SMOX), a member of the mammalian polyamine catabolic pathway, encodes an enzyme with cytoplasmic and nuclear expression in most tissues^42^, that catalyzes the oxidation of spermine to spermidine with the production of hydrogen peroxide (H_2_O_2_) and 3-aminopropanal (3-AP)^10–11^. SMOX has been reported as a source of induced reactive oxygen species (ROS) associated with neuroblastoma, gastric, lung, breast, prostate and colon cancers^12–16,36,37^, implying that inhibition of SMOX could be a target for chemoprevention^3^. However, so far, studies of SMOX inhibition have been observational in their nature with no direct inference on causation^12–16,36,37^. Genome-wide association studies (GWAS) of SMOX activity are also lacking. A GWAS could potentially detect genetic determinants of SMOX activity that might serve as instruments to assess the causality of SMOX on cancer using a Mendelian randomization framework^17^.

To our knowledge, there has not been any Mendelian randomization study assessing the potential causal relationship between SMOX activity and risk of any type of cancer. In this study, we present the first GWAS of spermidine/spermine ratio as a proxy for SMOX activity. We then use the genetic determinants of SMOX activity to perform Mendelian randomization analysis to assess causal relationships between SMOX activity and risk of lung, breast, prostate and colon cancer. In addition, we perform a phenome-wide association study (PheWAS) to query possible deleterious effect of altered SMOX activity through all disease categories and to test possible pleiotropic effects.

## METHODS

### Study population

The GWAS of spermidine/spermine ratio reflecting SMOX activity, was based on dried blood spot samples taken during routine newborn screening from 534 individuals of Danish ancestry, recruited in a matched case-control study of infantile hypertrophic pyloric stenosis (IHPS), as previously described^22,23^ (SSI-IHPS cohort). In addition, dried blood spot samples of 262 newborns of Danish ancestry were included for expression quantitative trait locus (eQTL) analysis (PSYCH-twin cohort). These 262 newborns were taken at random from each of 262 monozygotic twin pairs discordant for later diagnosis of psychiatric disorders as previously described^34^. Of these 262 newborns, 235 were included for methylation quantitative trait loci (mQTL) analysis. Furthermore, 671 whole blood samples taken from adults of the Estonian Genome Center University of Tartu (EGCUT) cohort^46^ were also used for eQTL (N=508)^35,47^ and mQTL (N=305; overlap with gene expression dataset 141 samples) lookups. The Danish Scientific Ethics Committee, the Danish Data Protection Agency and the Danish Neonatal Screening Biobank Steering Committee approved the PSYCH-twins study as well as the GWAS of metabolites in newborns. Usage of EGCUT RNA-seq dataset was approved by Estonian Committee on Bioethics and Human Research, protocol nr. 1.1-12/624, data extraction nr. N26.

The study also included GWAS summary statistics from four cancer-specific consortia cohorts: The ELLIPSE consortium (prostate cancer)^18^ with 79,194 cases and 61,112 controls; the BCAC consortium (breast cancer)^19^ with 62,533 cases and 60,976 controls; the TRICL consortium (lung cancer)^20^ with 29,266 cases and 56,450 controls, and the North American-based Children’s Oncology Group (neuroblastoma)^38^ with 2,101 cases and 4,202 controls. GWAS summary statistics for gastric and colorectal cancers relied on population-based cohorts: UK Biobank with 5,693 colorectal cancer cases and 386,740 controls^21^, and the BioBank Japan Project with 6,563 gastric cancer cases and 195,745 controls^33^. Analysis were restricted to individuals of European ancestry, except for gastric cancer due to the lack of publicly available European gastric cancer GWAS data.

Patients or the public WERE NOT involved in the design, or conduct, or reporting, or dissemination plans of our research.

### Metabolite measurements and GWAS

The metabolites spermidine and spermine were quantified in whole blood of dried blood spots using the AbsoluteIDQ® p400 Kit (Biocrates Life Sciences AG, Innsbruck, Austria). The AbsoluteIDQ® p400 Kit allows quantification of 408 metabolites which were described in our previous study^23^. In our previous study^23^, we quantified 148 of the 408 metabolites, from which spermidine and spermine measurements were also used for this study. In this study we performed genome-wide association scans of all these 148 metabolites and their biologically relevant ratios (e.g. direct substrate and product of an enzyme), including spermidine to spermine ratio, which had the most significant genome-wide hit of all the metabolites and ratios tested. For accuracy and computing time issues, the GWAS analysis was first implemented in PLINK^41^ and then repeated in R^24^ only for the genome-wide significant variants. The PLINK^41^ and R^24^ analyses were done using the following linear model: spermidine/spermine inverse normally transformed concentration ratios ∼ SNP + sex + YOB (as factor) + GA + Parity + IHPS. YOB pertains to year of birth, GA as gestational age in weeks, and IHPS as case/control status. As described before^22^, the SSI-IHPS cohort was array-genotyped with the Illumina Multi-Ethnic Global_v2_A2 array, and after genotyping QC, unobserved genotypes were imputed from the Haplotype Reference Consortium panel^43^. Altogether, 6,846,507 SNPs with minor allele frequency>=1% and imputation info score>=0.8 were used in the analysis.

### eQTL and mQTL lookups

To replicate the genetic association with SMOX activity at the gene expression level, eQTL analysis was performed in whole blood from two additional data sets of 262 newborns (a subset of the PSYCH-twin cohort) and 508 adults (a subset of the EGCUT cohort) that had previously been RNA-sequenced and GWAS-genotyped^34,35,47^. Preprocessing of the EGCUT expression data is described elsewhere^35^. Briefly, in EGCUT, gene expression matrix was normalized by trimmed mean of M values (TMM)^48^, log2 transformed, gene expression values were centered and scaled, and genotype multidimensional scaling (MDS) was regressed out. In addition, in order to remove non-genetic variance from the data, up to 20 gene expression-based PCs which were not associated with genetics were regressed out from the residuals of the expression data. Finally, inverse normal transformation was applied on the residuals from the previous step. Moreover, 235 out of 262 individuals from the newborn PSYCH-twin cohort^34^ also had genome-wide methylation array data from the Infinium HumanMethylation450 BeadChip (450k) array, as previously described^34^. For this study, all samples were filtered by Call Rate>0.99 (for detection P<0.01 and bead count >2) and median methylation and unmethylation intensity signal >2000 to ensure proper sample measurement quality. All probes (CpGs) were filtered by Call Frequency>0.99 (for detection P<0.01 and bead count >2) to ensure sufficient probe representation across most samples. The data was normalized with the noob background dye correction method after sample filtering^39^, implemented in the R^24^ package minfi. For all the 480 CpG sites within 1 Mb of the *SMOX* gene, we tested their methylation fraction against the lead (most significant) *SMOX* SNP. The analysis was implemented in R^24^ as the following linear model: CpG methylation fraction ∼ lead *SMOX* SNP + sex + YOB + GA + BW (birth weight). An association was deemed significant if P-value<0.05/480= 1.04×10^−4^ to correct for the number of CpGs tested. Furthermore, 305 individuals from the adult EGCUT cohort also had genome-wide methylation array data from the Infinium HumanMethylation450 BeadChip (450k) array. Briefly, methylation data was normalized according to the GoDMC (http://www.godmc.org.uk/) pipeline^44^ using functional normalization method as implemented in the R package meffil^45^. In step 1, we adjusted normalized methylation proportion for age, sex, predicted cell counts, predicted smoking and genetic PCs. In step 2, PCA was applied on the residuals from the previous step, and top non-genetic PCs were regressed out. To be able to estimate the SNPxAge effect on methylation, inverse normally transformed methylation proportions were regressed against covariates from step 1 and methylation PCs from step 2.

### Variance explained, Mendelian randomization, and PheWAS analyses

The lead SNP located on chromosome 20q12 (in cis with *SMOX*) being genome-wide significantly (P<5×10^−8^) associated with spermidine/spermine ratio was used as the genetic instrument in the Mendelian randomization analyses. Variance explained by the genetic determinant of SMOX activity was calculated as the adjusted R^2^ of lm function in R^24^ implemented as spermidine/spermine inverse normally transformed concentrations ratio ∼ SNP, or normalized and inverse normally transformed gene expression value of *SMOX* ∼ SNP. The inverse-variance weighted method^25^, implemented in the R^24^ package MendelianRandomisation (v0.3.0) was used to test whether the association between SMOX genotype and incident cancers could be caused by differences in SMOX activity due to genotype. The primary analysis measured the association between genetically determined SMOX activity and the risk of neuroblastoma, gastric, lung, breast, prostate and colorectal cancers. The PhenoScanner bioinformatic tool^27^, a curated database of publicly available results from large-scale genetic association studies, was used for the PheWAS analysis and detect possible concurrent risk factors of the investigated cancer types (pleiotropy). The PhenoScanner database was queried from the MendelianRandomization R^23^ package. In our main analyses we tested six key outcomes, namely neuroblastoma, gastric, lung, breast, prostate and colorectal cancers. A P-value of less than 0.05/6 (number of cancers tested) = 0.008 was considered statistically significant.

## RESULTS

### Participant characteristics

A total of 534 newborn participants (SSI-IHPS cohort) were included in the GWAS of spermidine/spermine ratio to find the genetic instrument of SMOX activity (Methods). An additional 262 newborns (PSYCH-twin cohort) and 508 adults (EGCUT cohort) were included to confirm the genetic instrument of SMOX activity at the gene expression level (Table 1 and Methods). A total of 950,575 participants, including 79,194 participants who had pancreatic cancer, 62,533 with breast cancer, 29,266 with lung cancer, 5,693 with colorectal cancer, 6,563 with gastric cancer and 2,101 with neuroblastoma were included as the cancer outcome cohorts (Table 2 and Methods). Demographic characteristics of each cohort are described in the original studies^18–23,33–35^.

**Table 1.** SNP rs1741315 associations with SMOX activity, *SMOX* gene expression and cg07472708 methylation fraction. SMOX activity and *SMOX* gene expression estimates were based on inverse normally transformed values (Methods). SNP rs1741315 ‘A’ allele is the effect allele. Pos, genomic position of rs1741315 (hg19); rsID, variant identifier; EAF, effect allele frequency; N, sample size; Beta, effect estimate; SE, standard error; R^2^, variance explained of rs1741315 on SMOX activity, *SMOX* gene expression and cg07472708 methylation fraction.

| Pos | rsID | Cohort | Analysis | N | EAF | Beta | SE | P Value | R<sup>2</sup> |
| --- | --- | --- | --- | --- | --- | --- | --- | --- | --- |
| 20:4155948 | rs1741315 | Newborns (SSI-IHPS) | SMOX activity | 534 | 0.458 | -0.757 | 0.046 | 1.34x10<sup>-49</sup> | 0.318 |
|  |  | Newborns (PSYCH-twins) | *SMOX* expr. | 262 | 0.471 | -0.830 | 0.062 | 2.47x10<sup>-30</sup> | 0.366 |
|  |  |  | cg07472708 | 235 |  | -0.012 | 0.003 | 4.63x10<sup>-05</sup> | 0.005 |
|  |  | Adults (EGCUT) | *SMOX* expr. | 508 | 0.452 | -0.366 | 0.065 | 2.75x10<sup>-08</sup> | 0.057 |
|  |  |  | cg07472708 | 305 |  | -0.062 | 0.069 | 0.371 | 0.002 |

**Table 2.** Associations between genetically predicted SMOX activity, based on SNP rs1741315, and six site-specific cancers (prostate, breast, lung, colorectal and neuroblastoma). All estimations were based on the inverse-variance weighted method with fixed-effects, with A being the effect allele. EAF, effect allele frequency.

| Outcome | Cases, No. | Controls, No. | EAF | Odds Ratio (95% CI) | P Value |
| --- | --- | --- | --- | --- | --- |
| Prostate cancer | 79,194 | 61,112 | 0.430 | 0.98 (0.96-1.00) | 0.046 |
| Breast cancer | 62,533 | 60,976 | 0.434 | 0.99 (0.96-1.02) | 0.465 |
| Lung cancer | 29,266 | 56,450 | 0.442 | 0.97 (0.94-1.00) | 0.084 |
| Colorectal cancer | 5,693 | 386,740 | 0.448 | 1.03 (0.99-1.07) | 0.096 |
| Gastric cancer | 6,563 | 195,745 | 0.361 | 0.99 (0.95-1.03) | 0.543 |
| Neuroblastoma | 2,101 | 4,202 | 0.412 | 0.95 (0.88-1.03) | 0.182 |

### Genetic associations reflecting SMOX activity

We performed a GWAS of spermidine/spermine ratio, with two loci reaching genome-wide significance (P<5×10^−8^) (Figure 1, Methods). One locus was located on chromosome 11p13, having four intergenic SNPs in perfect LD (r^2^=1) with each other, namely rs3891324, rs7124637, rs2067666 and rs11031804 (P=1.50×10^−8^). These SNPs explained 5% of the variance in the spermidine/spermine ratio. The other locus was on chromosome 20q12, with the most significant SNP rs1741315 (P=1.34×10^−49^) being an intronic variant in the *SMOX* gene (Figure 2). This lead SNP explained 32% of the variance in the spermidine/spermine ratio (Figure 3, Table 1). Despite that the locus on chromosome 11p13 reaches genome-wide significance, it is located in trans regarding *SMOX*. Since trans-acting SNPs can often be pleiotropic (violating one of the principles of Mendelian randomization)^26^, and since their variance explained is relatively small (5%) compared to the top cis-acting variant rs1741315 (32%), we chose to use only the latter as a genetic instrument of SMOX for the Mendelian randomization analysis. To assess effects of rs1741315 on SMOX activity at the gene expression level, we performed eQTL analyses in two additional cohorts. In a newborn cohort of 262 individuals (PSYCH-twin)^34^ rs1741315 was associated with *SMOX* gene expression (P=2.47×10^−30^) explaining 37% of the variance (Table 1). In a cohort of 508 adults (EGCUT)^35^, rs1741315 was also associated with *SMOX* gene expression (eQTL P=2.75×10^−8^) explaining 6% of the variance (Table 1, eFigure 1). Of note, rs1741315 was array-genotyped in all cohorts, in contrast to being just imputed. We stratified the adult cohort (EGCUT) into different age groups (10-years intervals) in order to detect a possible age-dependent eQTL effect in early vs. late adulthood (eFigure 2), but found no significant interaction between donor age at blood draw and rs1741315 genotype (P=0.50) on *SMOX* expression (eFigure 3). Due to some missing values in the data, this age-stratified analysis was performed on 494 out of the total 508 adults. Moreover, 235 out of 262 individuals from the newborn PSYCH-twin cohort^34^ had genome-wide methylation array data (Methods). Zooming in on CpGs at the SMOX locus, we found rs1741315 to be associated with the methylation proportion of the CpG cg07472708 (mQTL P=4.63×10^−5^) explaining 0.5% of the variance in methylation proportion for this CpG (Table 1). Contrary to the newborn data, we did not find rs1741315 to be associated with the methylation proportion of the CpG cg07472708 (mQTL P=0.371) in 305 individuals from the adult EGCUT cohort who also had genome-wide methylation array data (eFigure 4, Table 1 and Methods). Similar to the eQTL data, we also stratified the adult cohort (EGCUT) into different age groups (10-years intervals) in order to detect a possible age-dependent mQTL effect (eFigure 5), but found no significant interaction between donor age at blood draw and rs1741315 genotype (P=0.48) on cg07472708 methylation levels (eFigure 6). The CpG cg07472708 is located in the same SMOX intron as rs1741315 and is under a region of transcription factor binding site for the transcription factor GATA-1 (Methods). It should be noted that the 50-mer probe containing cg07472708 overlaps with rs1741317 (37 bp apart from the C nucleotide of cg07472708), a high LD (r^2^=0.996) SNP with our lead rs1741315. This could lead to a technical artifact from poorer measurement conditions in those individuals carrying the minor allele, so the results from mQTL analysis should be interpreted with caution.

**Figure 1.**
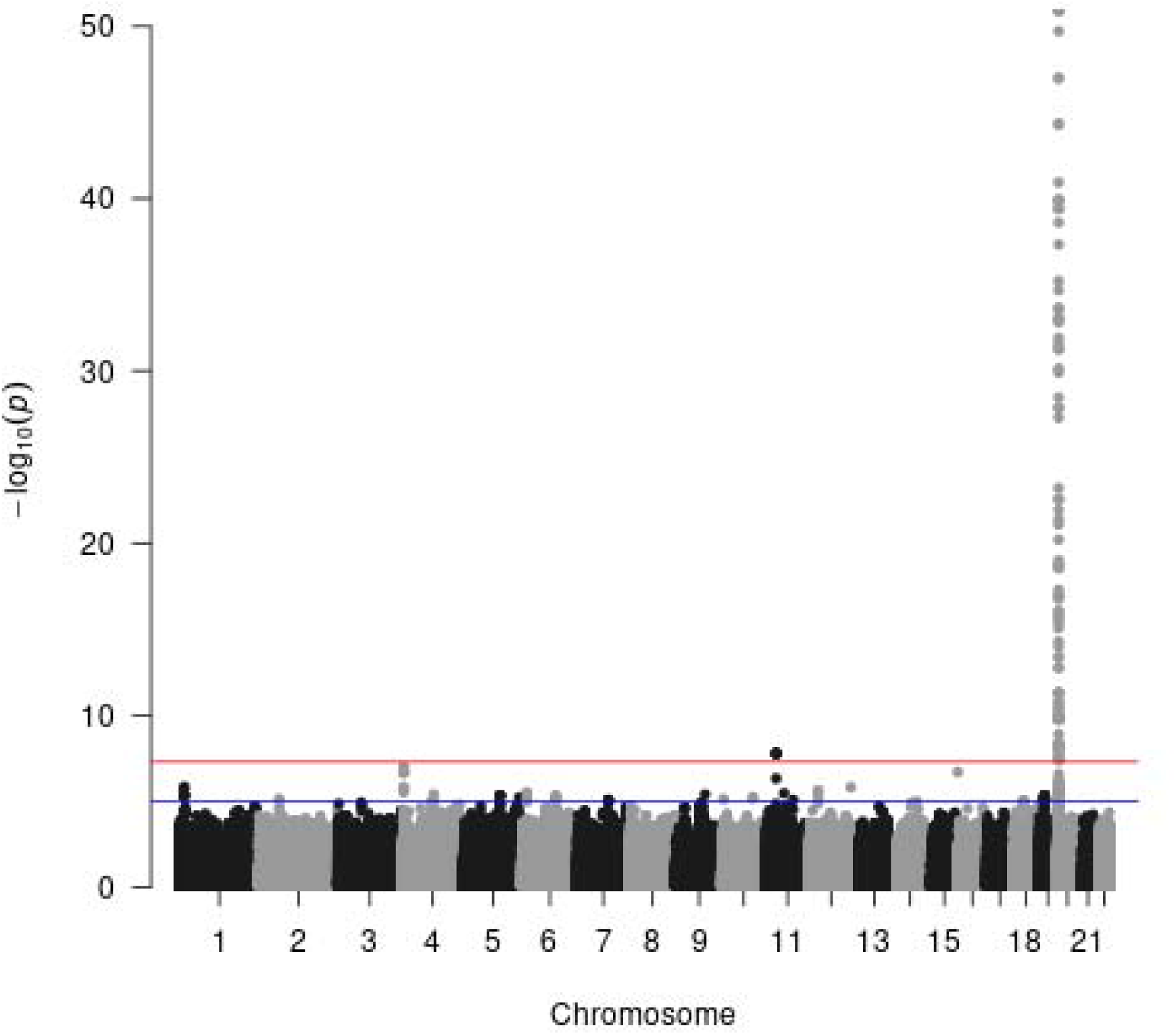
Genetic determinants of SMOX activity. Manhattan plot of the genome-wide association study (GWAS) of spermidine/spermine ratio (534 samples; SSI-IHPS cohort).

**Figure 2.**
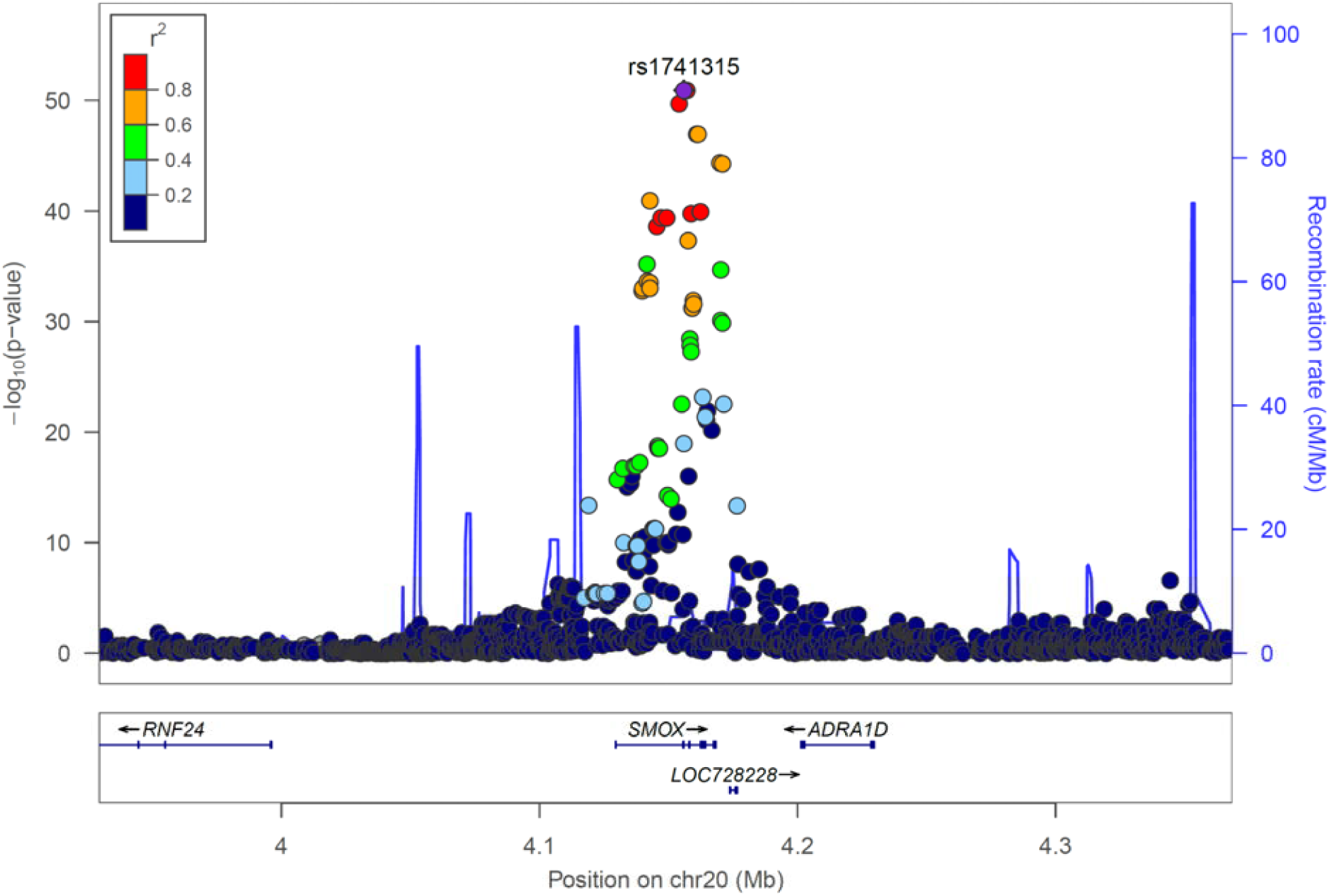
Regional association plot of the new genome-wide significant loci for spermidine/spermine ratio at the SMOX locus. Color-coded linkage disequilibrium (LD) is shown for the top SNP rs1741315 (LD determined with 1000Genomes EUR population). The x-axis represents the genomic region (hg19 assembly) surrounding 200 kb of SMOX gene, while the y-axis represents the strength of the association in −log10(P-value).

**Figure 3.**
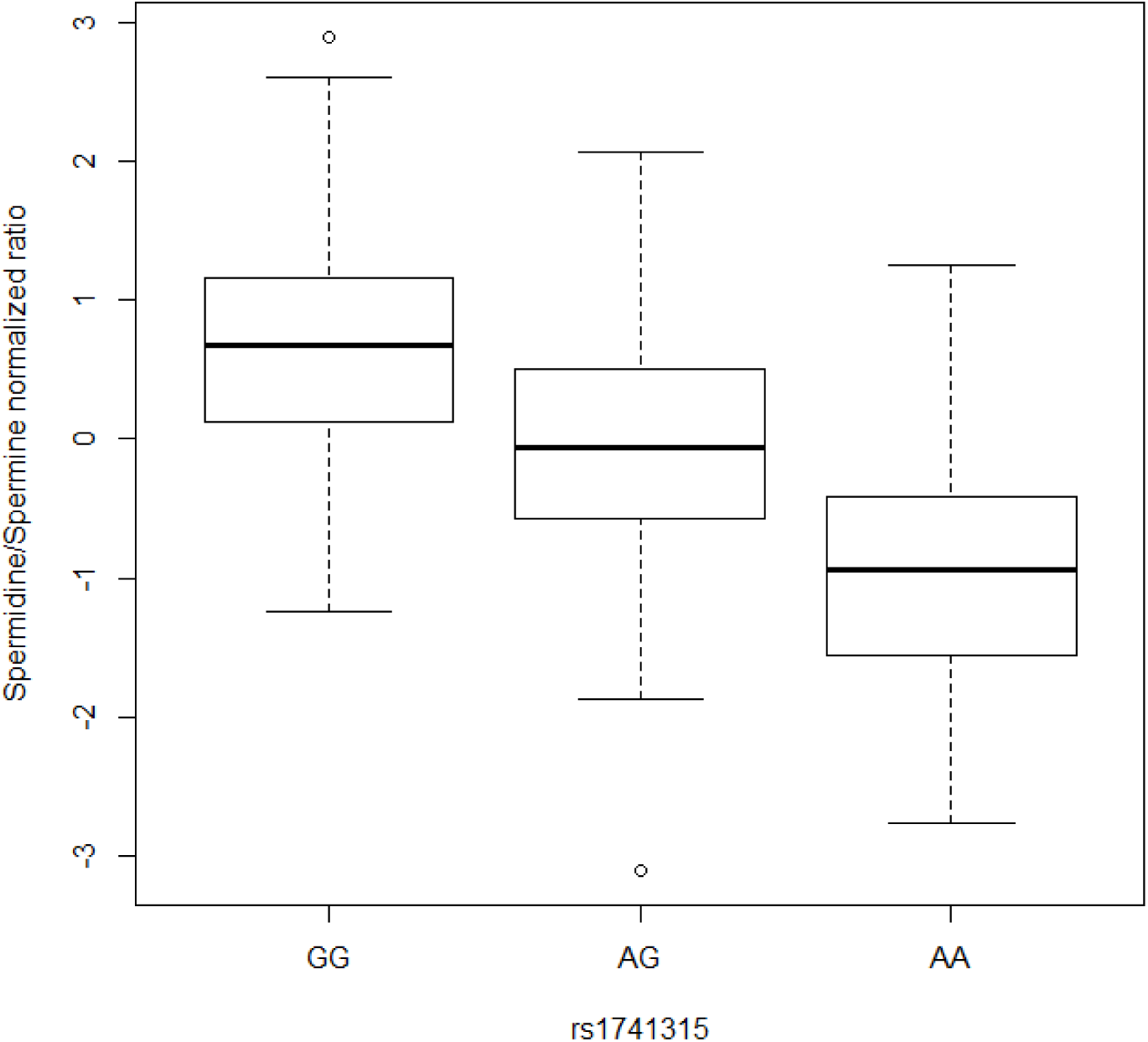
Boxplot of the rs1741315 alleles versus spermidine/spermine ratio (534 samples; SSI-IHPS cohort).

### PheWAS association of SMOX genetic instrument

One of the assumptions of Mendelian randomization is that genetic variants should only affect the outcome through their effect on the risk factors, i.e. no pleiotropic effects^29–30^. To test if this assumption holds, we checked the association of the SMOX genetic instrument rs1741315 against the PhenoScanner database of genotype-phenotype associations^27^ at P<1×10^−5^ (to correct for the 2998 traits/diseases tested in the database). With rs1741315 ‘A’ allele being the effect allele, we detected reticulocyte count (β=-0.150; 95% CI: −0.157, −0.143; P=4.12×10^−285^), lymphocyte count (β=0.016; 95% CI: 0.009, 0.023; P= 6.44×10^−6^) and hemoglobin concentration (β=0.016; 95% CI: 0.009, 0.023; P=8.05×10^−6^) as traits associated with this variant^31^. One potential way to test and adjust for pleiotropic effects through associations with these blood traits would be by multivariable Mendelian randomization analysis^26^. However, such analysis is only possible for multi-SNP genetic instruments, which we did not have.

### Association of SMOX genetic instrument with cancer risk

We found little evidence that the genetic instrument of SMOX activity, rs1741315, was associated with any of the five non-pediatric cancers evaluated (Table 2). Although genetically lower levels of SMOX were associated with slightly lower risk of prostate cancer at P=0.047, this finding should be interpreted with caution as it becomes non-significant when correcting for the six different cancers tested.

Since the SMOX genetic instrument, rs1741315, explained 5 to 6 times more variance of SMOX activity in newborns than in adults, we also tested the association between rs1741315 and neuroblastoma, a pediatric cancer in which SMOX has been reported to play a role^36,37^. Genetically lower levels of SMOX did not associate with lower risk of developing neuroblastoma^38^ (OR=0.95; 95% CI:0.88, 1.03; P=0.182) (Table 2).

## DISCUSSION

We conducted the first GWAS of spermidine/spermine ratio to identify genetic variants regulating spermine oxidase (SMOX) activity. Variants in the *SMOX* gene explained a large proportion of the variance in this ratio in newborns and were eQTLs for *SMOX* expression in newborns as well as adults. We did not find genetically determined SMOX activity to be associated with risk of either pediatric (neuroblastoma) or adult cancers (gastric, lung, breast, prostate, and colorectal cancer).

Observational studies have reported elevated SMOX levels in gastric, lung, breast, prostate, and colorectal cancers^12–16^, and it has been hypothesized that SMOX inhibition could be an effective target for chemoprevention^3^. Our results do not indicate that genetically driven variation in SMOX activity plays a major role in cancer risk. Several factors might explain this discrepancy between the observational epidemiological evidence^12–16^ and our Mendelian randomization results. Our results indicate that the genetic instrumental variable more accurately reflects SMOX activity in infancy, compared with later life, where most cancers occur. The positive association between observed SMOX levels and cancer in observational studies might be driven by reverse causation, in which cancer in an individual could cause SMOX to be elevated due to inflammation. In addition, elevated SMOX levels in cancer could also be due to environmental factors not captured by genetics, e.g. short-term pharmacological changes, induced by SMOX inhibitors. Our genetic instrument was developed based on normal range SMOX activity data, thus additional genetic variants might play a role in aberrant expression of this enzyme. Furthermore, despite we have corrected for disease status, half the samples from the discovery SSI-IHPS cohort were selected to be IHPS cases, and are therefore not representative of the general population. However, the association of rs1741315 with spermidine/spermine ratio did not differ between IHPS cases and controls (eFigure 7). The instrumental variable SNP is also associated with hematological traits^31^ that could potentially affect cancer risk. To assess whether pleiotropic effects drove our results, we performed a PheWAS analysis, with no diseases or non-hematological traits detected. We detected reticulocyte count, lymphocyte count and hemoglobin concentration associated with rs1741315. However, it is unclear if and how the variability in those blood cell phenotypes could affect the cancer predisposition. Therefore there was no reason to conclude that pleiotropy would affect our Mendelian Randomization analyses.

Our study had important strengths. Several previous studies which examined SMOX activity and cancer risk were susceptible to recall bias, confounding and reverse causation^11–17^ none of which are concerns of Mendelian randomization studies^17^. In addition, the fact that our genetic instrument explained a sizeable proportion of the variance of SMOX activity together with the fact that we used summary statistics from the largest meta-analyses of primary GWAS of these cancer types to date^18–21,33,38^ are factors enhancing the statistical power to detect causal effects^32^.

In conclusion, lifelong genetic exposure to low levels of SMOX was not associated with lower cancer risk. However, this finding cannot be assumed to represent a short-term strong drug inhibition of SMOX, thus establishing causality would require a randomized clinical trial. To better interpret the complexity of the relationship between SMOX activity and cancer, future studies should effectively distinguish newborn versus adulthood SMOX activity and short versus long exposures. This study is the first to use Mendelian randomization to assess the possible benefits of SMOX inhibitors on cancer risk, paving the way for other polyamine catabolic pathway enzymes to be tested with the same methodology.

## Data Availability

Data available upon request.

## ACKNOWLEDGMENTS

This study received partial support from the Danish Medical Reseach council (DFF 4004-00512), the Novo Nordisk Foundation (NNF18OC0053228) and the Oak Foundation (OCAY-18-598). It was conducted using the Danish National Biobank resource, supported by the Novo Nordisk Foundation, grant number 2010-11-12 and 2009-07-28. We would also like to acknowledge iPSYCH and The Lundbeck Foundation for their QTL validation datasets. The iPSYCH (The Lundbeck Foundation Initiative for Integrative Psychiatric Research) team acknowledges funding from The Lundbeck Foundation (grant numbers R102-A9118 and R155–2014-1724). This research has also been conducted using the UK Biobank Resource (project ID 31063) available from the Pan-UKB team. This work has also been supported by the European Regional Development Fund and the programme Mobilitas Pluss (MOBTP108), SP1GI18045T, No. 2014-2020.4.01.15-0012 GENTRANSMED and 2014-2020.4.01.16-0125. This study was also funded by EU H2020 grant 692145, Estonian Research Council Grant PUT1660 and PRG687. Data analyzes with Estonian datasets were carried out in part in the High-Performance Computing Center of University of Tartu. Furthermore, we acknowledge the Genetic Associations and Mechanisms in Oncology (GAME-ON)/Elucidating Loci Involved in Prostate Cancer Susceptibility (ELLIPSE) Consortium, the Transdisciplinary Research Into Cancer of the Lung (TRICL) consortium, the Breast Cancer Association Consortium (BCAC), and the North American-based Children’s Oncology Group. We further thank all the participants, staff and researchers of the Danish National Biobank, Estonian, Japan and UK biobanks, along with the cancer case-control cohorts above described for their contribution to this research.

