## Supplemental Figures S1-S7 for "Mendelian randomization study of spermine oxidase and cancer risk"

**eFigure 1.** Association between rs1741315 alleles and *SMOX* RNA-seq expression (508 adults; EGCUT cohort).

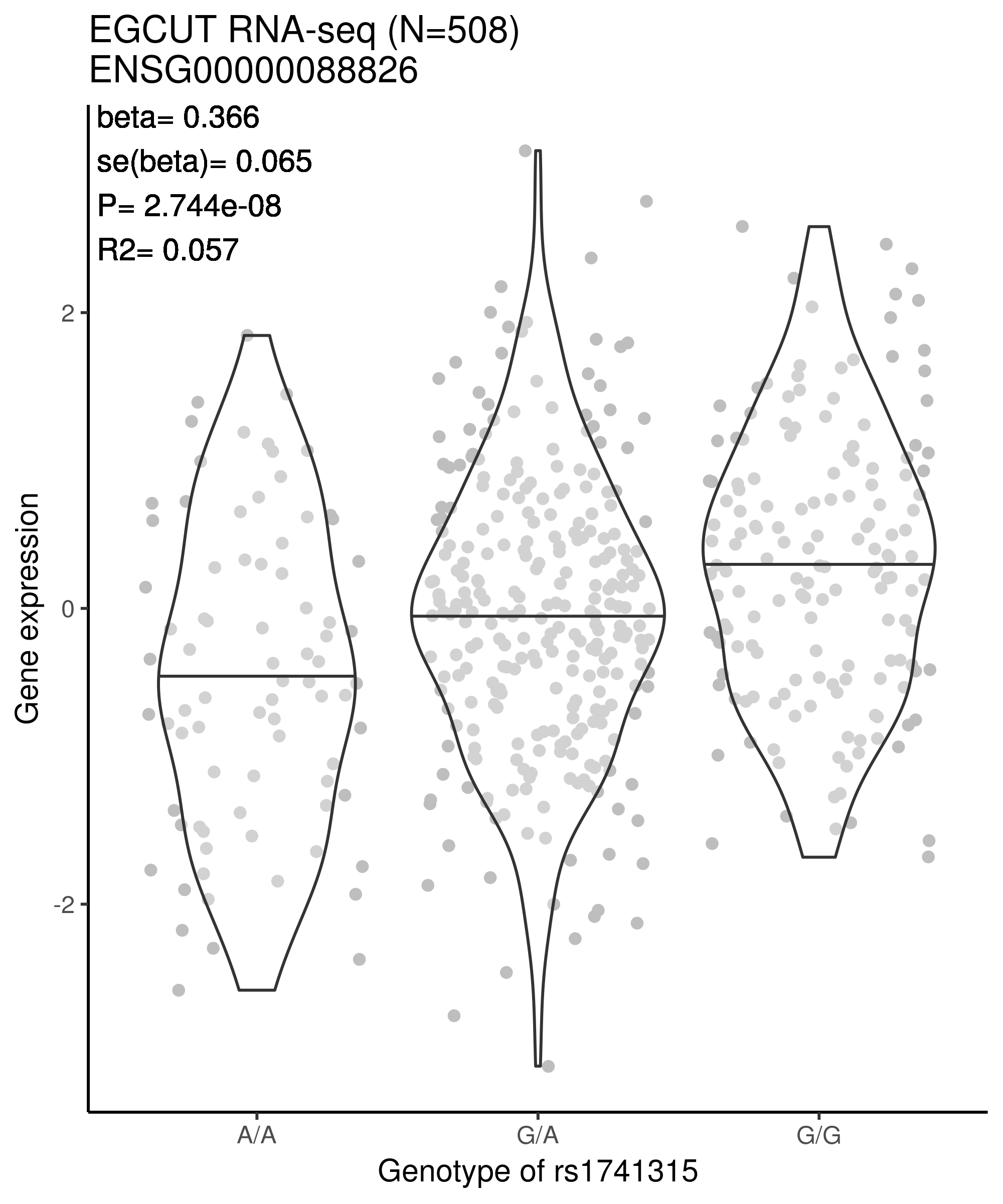

**eFigure 2.** Association between rs1741315 alleles and *SMOX* RNA-seq expression, stratified into 10-year age groups (494 adults; EGCUT cohort).

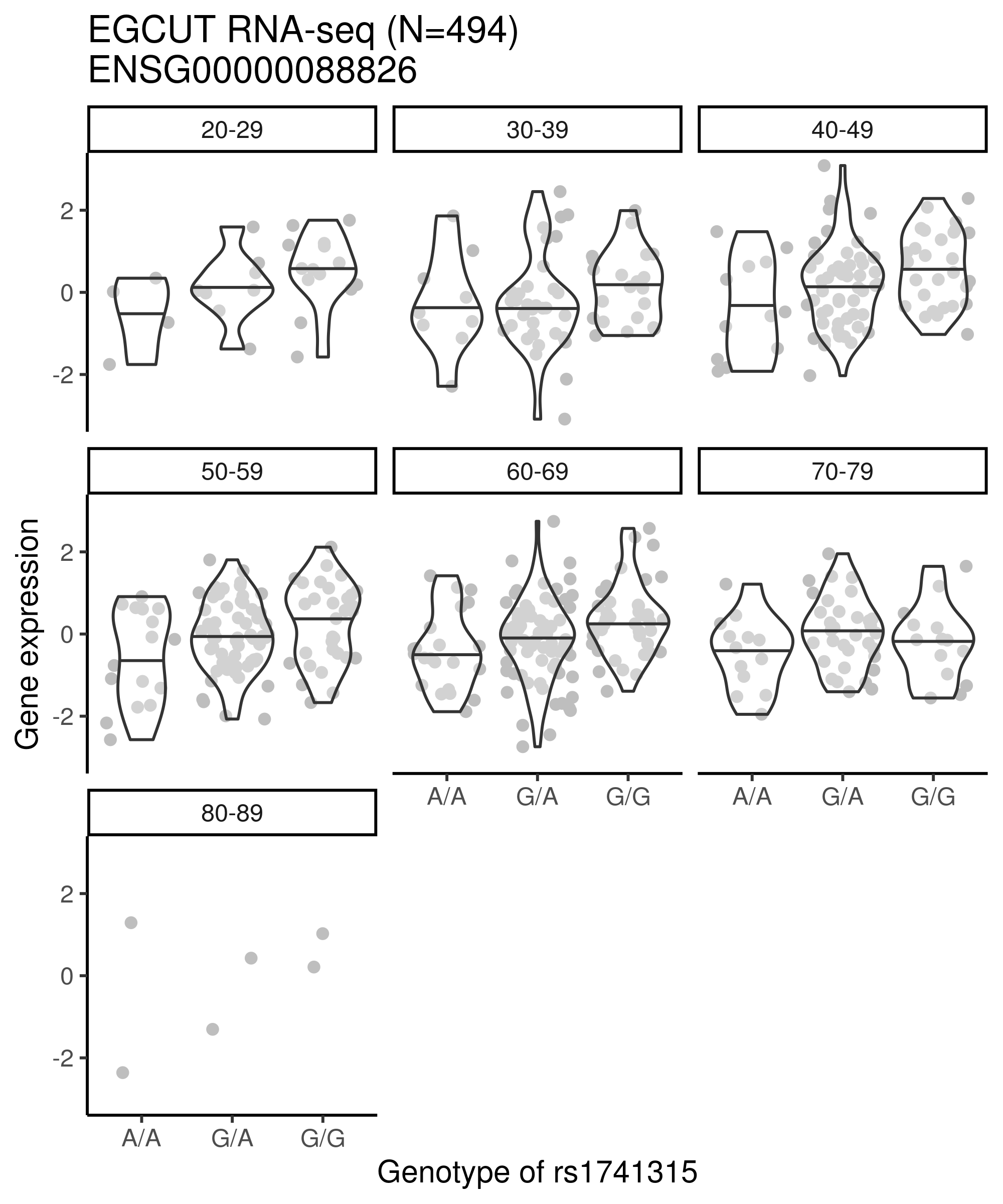

**eFigure 3.** Interaction between donor age at blood draw and rs1741315 genotype on *SMOX* expression (494 adults; EGCUT cohort).

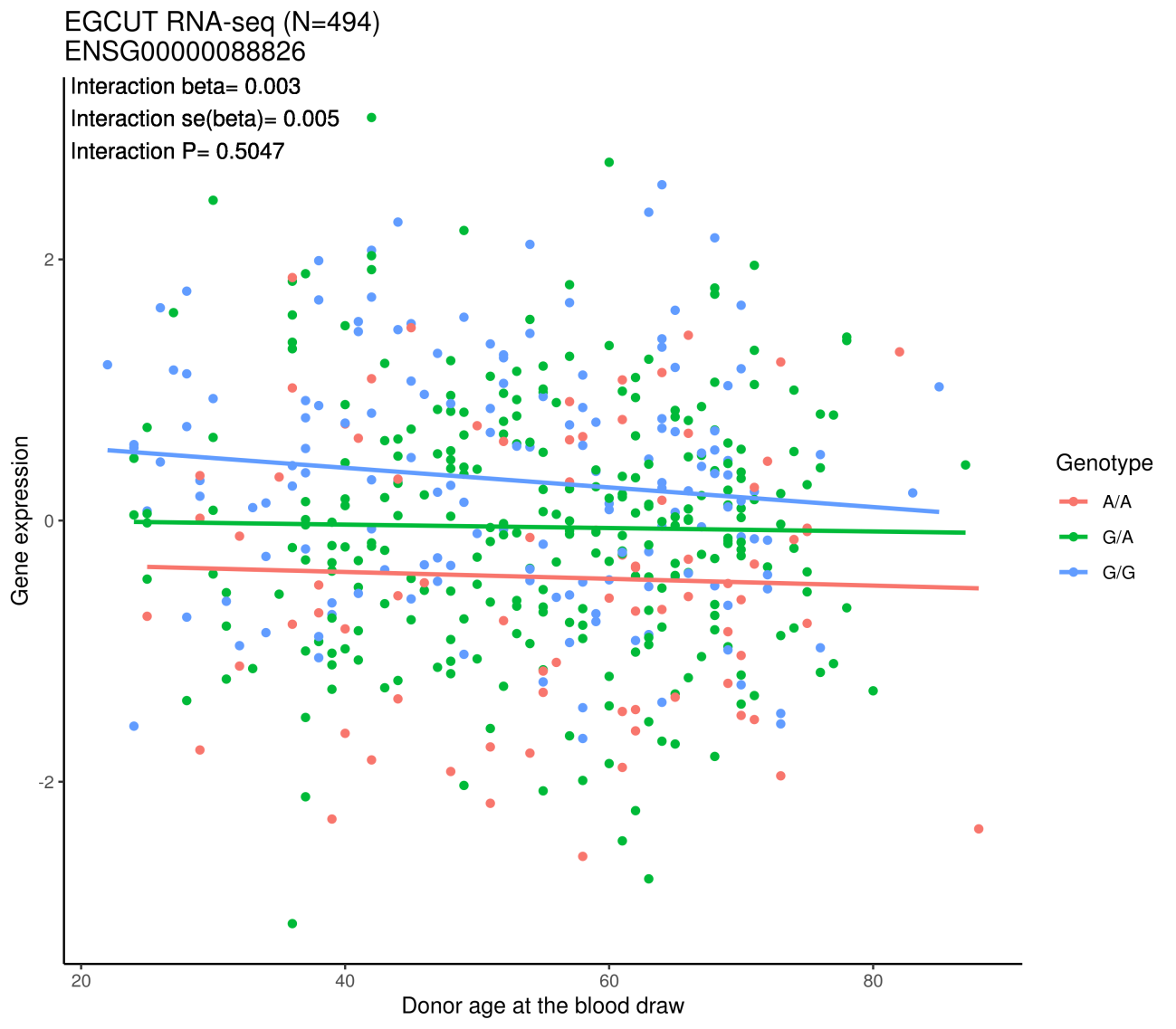

**eFigure 4.** Association between methylation proportion of the CpG cg07472708 and rs1741315 (305 adults; EGCUT cohort).

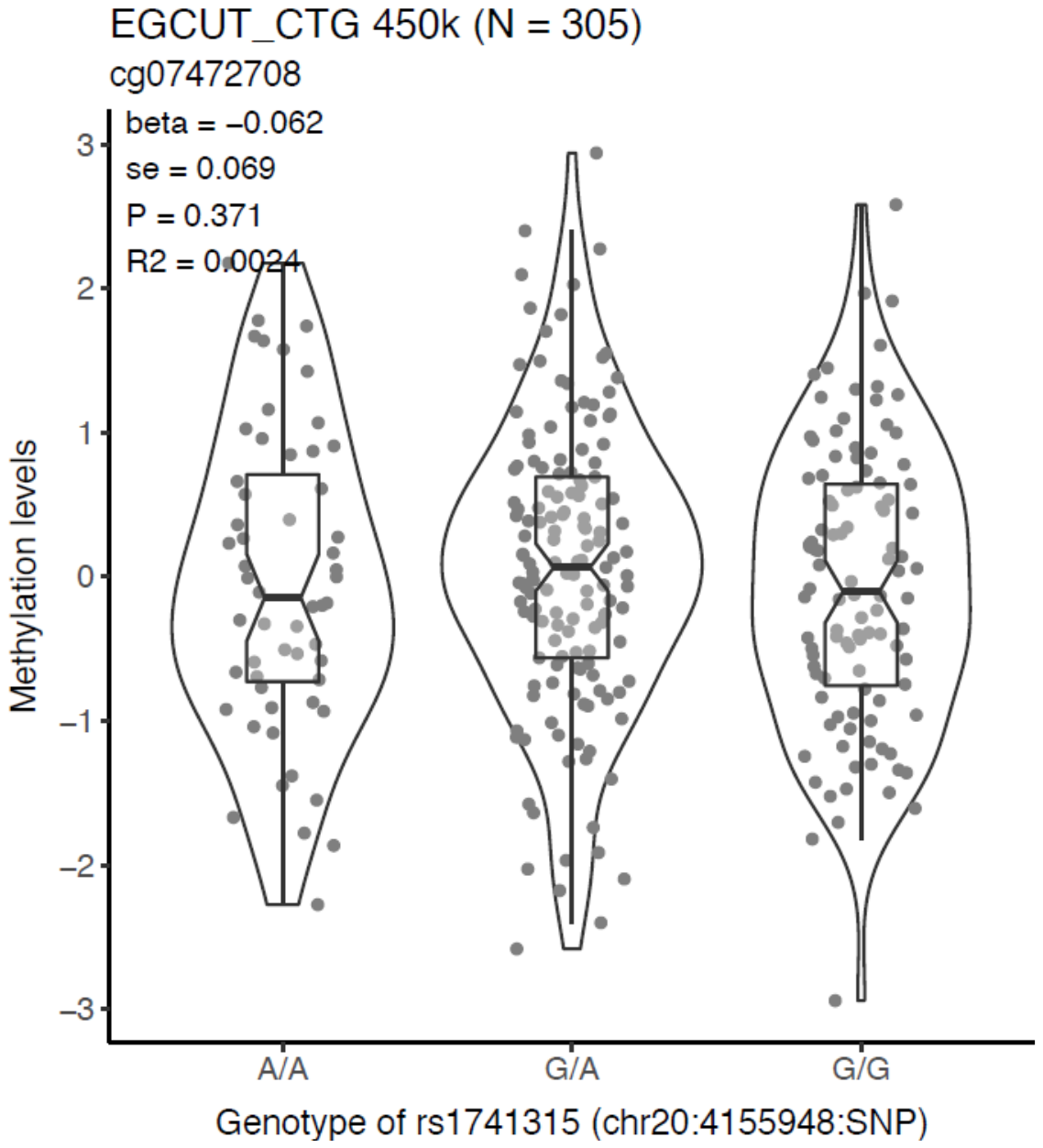

**eFigure 5.** Association between methylation proportion of the CpG cg07472708 and rs1741315, stratified into 10-year age groups (305 adults; EGCUT cohort).

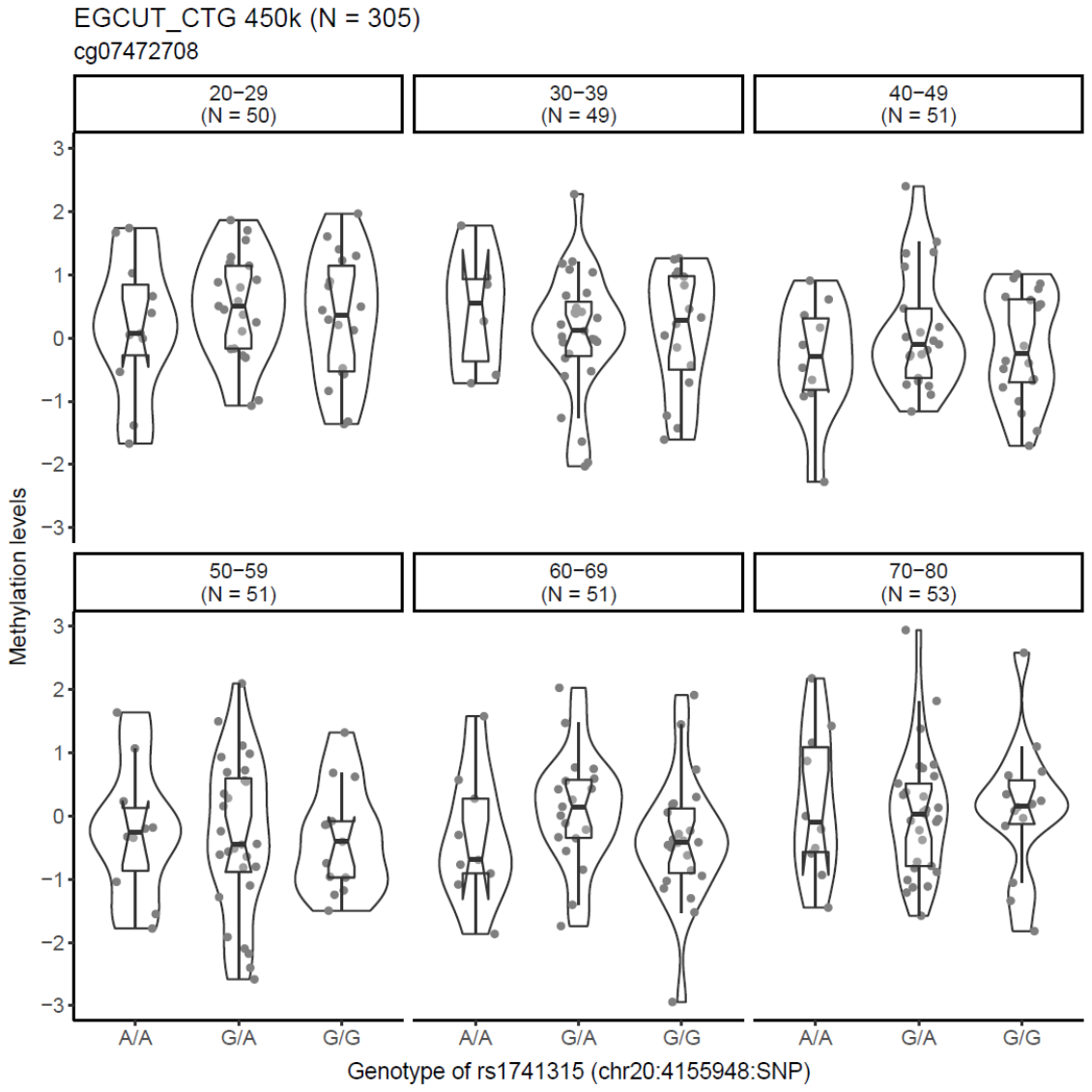

**eFigure 6.** Interaction between donor age at blood draw and rs1741315 genotype on cg07472708 methylation levels (305 adults; EGCUT cohort).

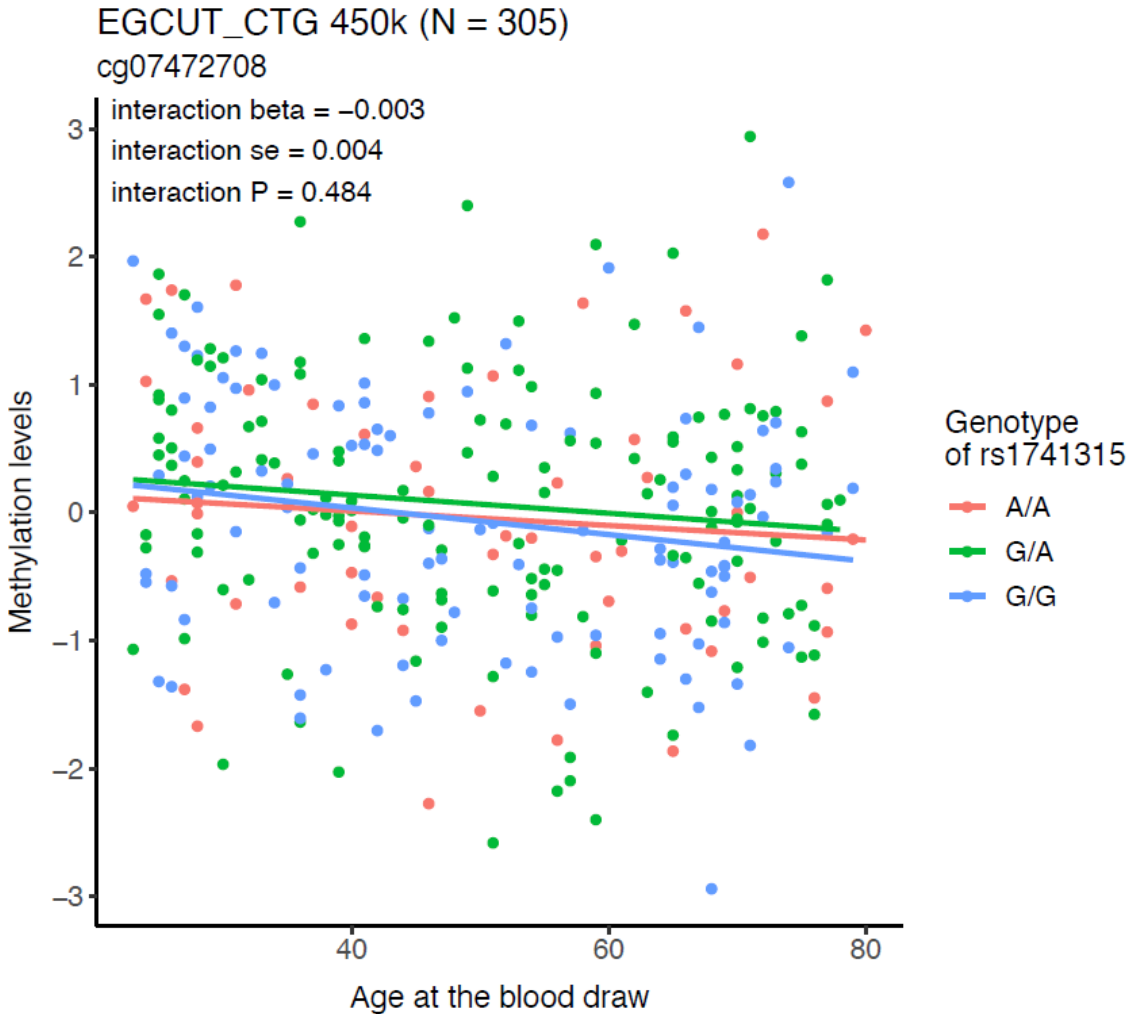

**eFigure 7.** Boxplot of the rs1741315 alleles versus spermidine/spermine ratio, stratified by IHPS disease status (534 samples; SSI-IHPS cohort). 1= control; 2 = IHPS case.

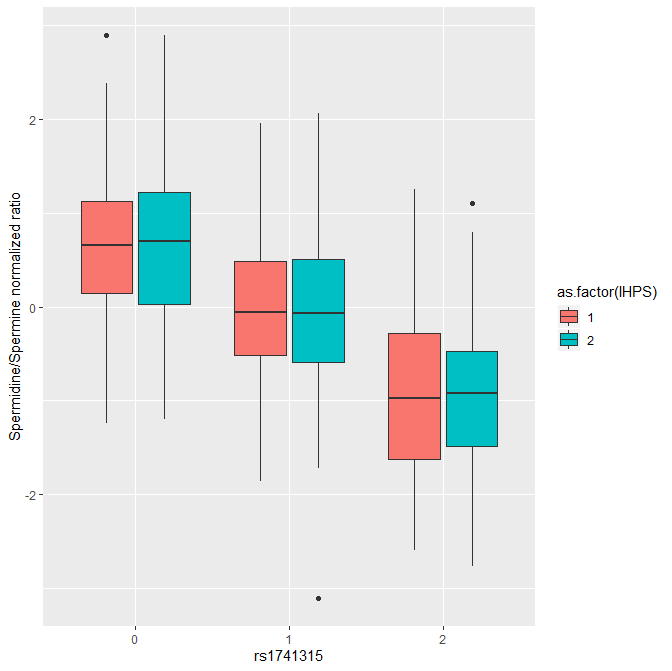
